# Low Covid-19 hospitalisation in Dumfries and Galloway: comparison with other Scottish health boards

**DOI:** 10.1101/2020.05.22.20110163

**Authors:** Andrew Rideout, Calum Murray, Chris Isles

## Abstract

**Background:** Covid-19 virus activity appears to have affected some parts of the United Kingdom more than others. Dumfries and Galloway (D&G) has seen fewer hospitalised cases than predicted. We wondered whether this might be related at least in part to population density.

**Methods:** We compared Covid-19 hospitalisation rates/100,000 population in D&G with those of the other 10 mainland Scottish health boards. We chose two time points: 19th April which was the peak of the pandemic in Scotland and 15th May, seven and a half weeks after lockdown. We used chi square and odds ratios with 95% confidence intervals to test for differences in hospitalisation rates and Pearson’s correlation coefficient to examine the relation between hospitalisation rates and population density. Population density for each health board was provided by National Records of Scotland.

**Results:** Hospitalisation in D&G was 13.4/100,000 on 19th April, falling to 1.3/100,000 by 15th May. Corresponding hospitalisation rates in Greater Glasgow & Clyde (GGC) were 50.1/100,000 and 38.9/100,000. Compared to GGC, hospitalisation rates in D&G were 3 times lower at peak (OR 0.27, 95% CI 0.17, 0.42) and 30 times lower by 15th May (OR 0.03, 95% CI 0.01, 0.14). Hospitalisation rates for the other health boards lay in between values recorded for D&G and GGC and fell in 10 of the 11 boards between these two dates. There was a positive association between hospitalisation rate and population density (r=0.756, p=0.007 on 19th April and r=0.840, p<0.001 for 15th May).

**Conclusion:** We have confirmed there are large differences in Covid-19 hospitalisation rates across the 11 mainland Scottish health boards, that are in part related to population density. These data support a regional rather than one nation approach to easing Covid-19 restrictions.

## Introduction

‘Stay at home, protect the NHS, save lives’. The British public have taken this message to heart and to greater degree than anticipated with 71% saying they would now be nervous about leaving their homes even if businesses were allowed to reopen and travel restrictions were lifted^1^. There were fears initially that the NHS would be overwhelmed by Covid positive admissions and that our critical care units would run out of ventilators. This led to the rapid construction of nine Nightingale hospitals across the UK, all of which have since been stepped down. So, if the main thrust of staying at home was to protect the NHS then the slogan and the campaign can be judged to have been successful.

It remains to be seen what happens when restrictions are lifted. There is clearly anxiety here with predictions of a second surge in infection if lockdown is relaxed too soon^2^. While much has been made of the UK’s Covid death toll, which as of 15th May was second highest in the world after the USA and fourth highest in the world when expressed per million population after Spain, Italy and Belgium, little has been written about regional variations in coronavirus activity within the UK or the possibility that an earlier or faster return towards normal activity might be possible in those areas where virus activity is low.

It is against this background and the clinical impression we were seeing fewer Covid positive patients than predicted that we undertook a survey of hospitalised Covid positive patients in Scotland. We were particularly interested in the possible influence of population density on the numbers of hospitalised cases.

## Methods

Dumfries and Galloway is a mostly rural region that stretches 100 miles from Stranraer in the west to Langholm in the east. The most recent estimate of population is 148,860^4^. The region is served by two hospitals: Dumfries and Galloway Royal Infirmary (DGRI) is a 331 bed district general hospital in Dumfries and the Galloway Community Hospital is a 44 bed community hospital in Stranraer. DGRI has a 17 bed Critical Care Unit.

We used Scottish Government data from 26th March onwards^5^ to document the number of Covid positive patients in hospital at midnight each night per 100,000 population in each of the eleven Scottish mainland health boards. We chose two time points: 19th April which was the peak of the pandemic in Scotland and 15th May seven and a half weeks after lockdown. Our analysis was in two parts. First, we compared hospitalisation rates in Dumfries and Galloway with those of Ayrshire and Arran, a neighbouring health board; with Highland, the least densely populated health board; and with Greater Glasgow & Clyde, the largest and most densely populated board in Scotland. Second, we correlated hospitalisation rates across all mainland health boards with population density in each board^4^.

We chose not to analyse the number of Covid positive cases in each health board because of the risk of bias due to different testing regimens. Nor did we attempt to compare death rates. This was because some deaths recorded as due to Covid-19 are possible or presumed Covid in patients who have not tested positive for the virus (clinical diagnosis rather than laboratory diagnosis) and also because not everyone who has died with Covid symptoms has been tested for the virus. We compared hospitalisation rates between boards by chi square test and odds ratios with 95% confidence intervals where appropriate and correlated hospitalisation rates with population density using Pearson’s correlation coefficient.

## Results

### Daily hospitalisation rates in four mainland health boards

Figure 1 shows daily hospitalisation rates for Covid positive patients per 100,000 population in Dumfries and Galloway, Ayrshire and Arran, Highland, Greater Glasgow & Clyde and Scotland (shaded). Hospitalisation rates in Dumfries and Galloway have been lower than in the neighbouring health board of Ayrshire and Arran (OR 0.55, 95% CI 0.34, 0.88 on 19th April; OR 0.12, 95% CI 0.03, 0.49 on 15th May); similar to those recorded in Highland health board (OR 0.96, 95% CI 0.57, 1.63 on 19th April; OR 0.86, 95% CI 0.17, 4.46 on 15th May); and considerably less than in Greater Glasgow and Clyde. Compared to Greater Glasgow and Clyde, hospitalisation rates in Dumfries and Galloway were around 3 times lower on 19th April (OR 0.27, 95% CI 0.17, 0.42) and 30 times lower on 15th May (OR 0.03, 95% CI 0.01, 0.14).

**Figure 1.**
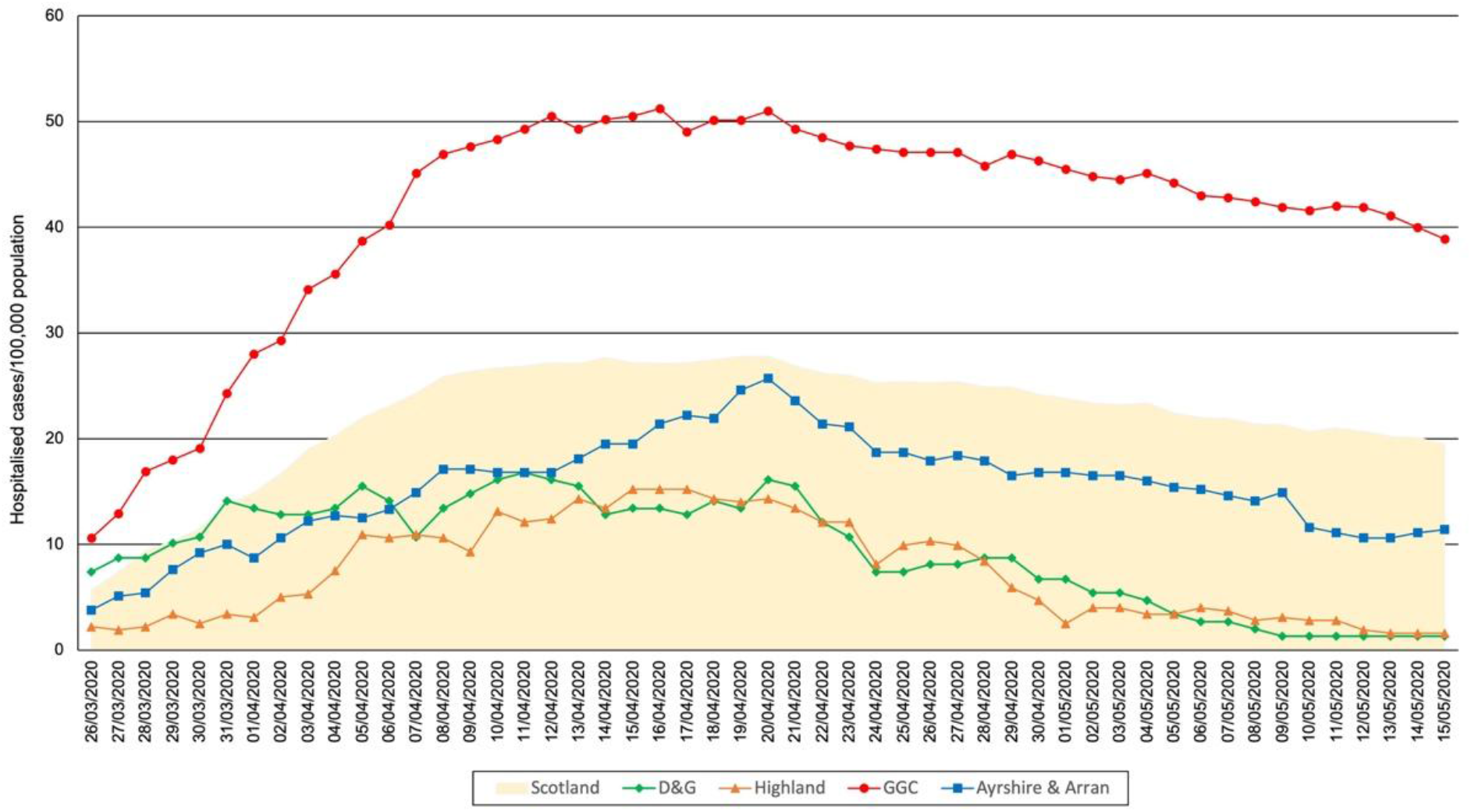
Daily hospitalisation rates for confirmed Covids/100,000 population in Dumfries and Galloway, Ayrshire & Arran, Highland, Greater Glasgow and Clyde and Scotland (shaded)

### Hospitalisation rates and population density

Table 1 shows hospitalisation rates for Covid positive patients per 100,000 population in each Scottish mainland health board on 19th April and 15th May. Hospitalisation rates fell in 10 of 11 health boards between these two dates. Three of the four health boards with the highest hospitalisation rates on both dates had population density greater than 200 persons/100,000 (the exception was Borders) while the four health boards with the lowest hospitalisation rates all had population density less than 150 persons/100,000 (Table 1). Statistical analyses confirmed a significant inter board variation in hospitalisation rates for 19th April (chi square for trend 268.4, p<0.001) and for 15th May (chi square for trend 350.0, p<0.001); and a positive association between hospitalisation rates and population density for the 11 mainland boards on both dates (r=0.756, p=0.007 for 19th April and r=0.840, p<0.001 for 15 th May).

**Table 1.** Population, population density and confirmed Covid-19 hospitalisation rates/100,000 population on 19th April and 15th May.

| Health Board | Population Mid June 2019 | Population density/ km<sup>2</sup> | 19<sup>th</sup> April Confirmed cases in hospital | Confirmed cases in hospital/ 100,000 (95% CI) | 15<sup>th</sup> May Confirmed cases in hospital | Confirmed cases in hospital/ 100,000 (95% CI) |
| --- | --- | --- | --- | --- | --- | --- |
| Ayrshire & Arran | 369,360 | 110 | 91 | 24.6 (19.6, 29.7) | 42 | 11.4 (7.9, 14.8) |
| Borders | 115,510 | 24 | 43 | 37.2 (26.1, 48.4) | 25 | 21.6 (13.2, 30.1) |
| Dumfries & Galloway | 148,860 | 23 | 20 | 13.4 (7.5, 19.3) | 2 | 1.3 (0.0, 3.2) |
| Fife | 373,550 | 282 | 105 | 28.1 (22.7, 33.5) | 75 | 20.1 (15.5, 24.6) |
| Forth Valley | 306,640 | 116 | 50 | 16.3 (11.8, 20.8) | 21 | 6.8 (3.9, 9.8) |
| Grampian | 585,700 | 67 | 72 | 12.3 (9.5, 15.1) | 96 | 16.4 (13.1, 19.7) |
| Greater Glasgow & Clyde | 1,183,120 | 1072 | 593 | 50.1 (46.1, 54.2) | 460 | 38.9 (35.3, 42.2) |
| Highland | 321,700 | 10 | 45 | 14.0 (9.9, 18.1) | 5 | 1.6 (0.2, 2.9) |
| Lanarkshire | 661,900 | 295 | 180 | 27.2 (23.2, 31.2) | 113 | 17.1 (14.9, 20.2) |
| Lothian | 907,580 | 526 | 228 | 25.1 (21.9, 28.4) | 200 | 22.0 (19.0, 25.1) |
| Tayside | 417,470 | 55 | 90 | 21.6 (17.0, 26.0) | 23 | 5.5 (3.3, 7.8) |
| Scotland | 5,463,300 | 70 | 1520 | 27.8 (26.4, 29.2) | 1066 | 19.5 (18.3, 20.7) |

## Discussion

The results of our survey show significantly lower hospitalisation rates for Covid positive patients in Dumfries and Galloway when compared to Greater Glasgow & Clyde and to Scotland as a whole. Hospitalisation peaked 2-3 weeks after lockdown and has been falling steadily since. At no point since the pandemic began has the NHS been overwhelmed in south west Scotland, suggesting that lockdown has successfully reduced the spread of the virus in this mostly rural region. Highland health board have reported similarly low hospitalisation rates.

So why has Dumfries & Galloway responded so well to lockdown? The number of Covid positive cases per 100,000 population in D&G and GGC on 23rd March were essentially the same at 12.1 and 12.8 respectively (OR 0.94, 95% CI 0.58, 1.53), suggesting that both health boards locked down at approximately the same stage of the pandemic. Could age^6^, ethnicity^7^ or population density have been responsible for lower hospitalisation rates? Dumfries and Galloway has the highest proportion of older adults in Scotland with 25% of the population aged 65 years or over^9^ while ethnic minorities accounted for only 1% of the population at the 2011 census^10^. Corresponding figures for Greater Glasgow & Clyde are 16.2% over 65 in 2013 and 7.5% ethnic minorities in 2011^11^. Taken together it seems unlikely that these differences could account for a 30 fold difference in hospitalisation rates. Differences in population density, on the other hand, seem an altogether more plausible explanation, if for no better reason than that a low population density must make it easier to practice social distancing. We recognise, however, that population density cannot be the only explanation given the relatively high rate of hospitalisation in neighbouring Borders health board (Table 1).

‘Guided by the science’ has become the mantra of the official response to anyone who questions its approach to the coronavirus crisis. Lockdown may well have prevented the NHS from being overwhelmed but has certainly been harmful in other respects: the economy remains shut down, the mental health of the nation is said to be suffering^12^ and domestic violence is on the increase^13^. The UK government has said there are five conditions that require to be met before coronavirus lockdown can be lifted. These are that there should be sufficient critical care capacity; a sustained and consistent fall in daily deaths; evidence that the rate of infection is decreasing; that testing and PPE are able to meet demand; and that any relaxation of measures will not risk a second peak that overwhelms the NHS. It is our belief that a regional approach to easing restrictions may be logical if backed by real time data and that if regions such as ours are meeting these five criteria already we should not have to wait for parts of the country with higher levels of virus activity to do so before beginning the slow return to normality^14^.

In conclusion, we have confirmed there are large differences in Covid-19 hospitalisation rates across the 10 mainland Scottish health boards, that are in part related to population density. These data support a regional rather than one nation approach to easing Covid-19 restrictions.

## Data Availability

All data available on request to corresponding author

## Declarations

### Contributorship

- CI had the idea for the project; CM compiled the hospitalisation data from the Scottish Government Covid website; AR was responsible for the statistical analyses; all authors contributed to and approved the final version.

### Data availability

– all data available on request to corresponding author

